# Genetic and Epigenetic Contributors to Cleft Laterality: Evidence from Monozygotic Mirror Twins and Replication Cohorts

**DOI:** 10.1101/2024.11.16.24317351

**Authors:** Aline L. Petrin, Waheed Awotoye, Christina E. Spencer, Ligiane A. Machado-Paula, Luke Hovey, Henry Keen, Michael Chimenti, Patrick Breheny, Fang Qian, Shareef Dabdoub, James C. Thomas, Azeez Butali, Jeffrey C. Murray, Shankar Rengasamy Venugopalan, Lina M. Moreno-Uribe

**Affiliations:** Department of Orthodontics, College of Dentistry and Dental Clinics, University of Iowa, Iowa City, IA, 52242, USA; College of Dentistry and Dental Clinics, University of Iowa, Iowa City, IA, 52242, USA; Iowa Institute of Human Genetics, Carver College of Medicine, University of Iowa, Iowa City, IA, 52242, USA; Department of Orthodontics, Tufts University School of Dental Medicine, Boston, MA, 02111, USA; Department of Biostatistics, College of Public Health, University of Iowa, Iowa City, IA, 52242, USA; Division of Biostatistics and Computational Biology, College of Dentistry and Dental Clinics, University of Iowa, Iowa City, IA, 52242, USA; Department of Oral Pathology, Radiology, & Medicine, College of Dentistry and Dental Clinics, University of Iowa, Iowa City, IA, 52242, USA; Department of Pediatrics, Carver College of Medicine, University of Iowa, Iowa City, IA, 52242, USA

## Abstract

Nonsyndromic cleft lip (nsCL) exhibits a non-random laterality pattern, with left-sided clefts occurring approximately twice as frequently as right-sided clefts. The molecular mechanisms underlying this laterality bias remain poorly understood.

We performed whole-genome sequencing and methylation profiling on a family comprising monozygotic twins with mirror-image nsCL, their affected mother, and unaffected father and brother. We conducted three independent replications via (1) publicly available whole genome data; (2) genome-wide methylation analysis in 38 individuals with unilateral cleft; and (3) validation of methylation results in the top 3 candidate genes in 385 unrelated individuals with unilateral clefts (DNA from blood or saliva).

We identified a variant in *FGF20* (p.Ile79Val) shared by the twins and their mother. We observed laterality and severity-associated methylation differences in three main genes. *ARID5B* showed higher methylation in left clefts (saliva, p=0.001; blood, p=0.032). *ZFP57* demonstrated a strong cleft-extent effect, with cleft lip and palate (CLP) showing markedly higher methylation than cleft lip only (CL) (LCLP vs. RCL p^adj^=0.0004; LCLP vs. LCL p^adj^ = 0.019). *HOOK2* displayed a cross-tissue cleft-extent effect in the opposite direction — CLP subtypes were hypomethylated relative to CL-only subtypes in blood (p<0.0001) and saliva p=0.0008).

This study provides evidence that DNA methylation patterns plays a role in both the laterality and severity of cleft lip. *ARID5B* provides a consistent laterality signal across tissues, while *ZFP57* and *HOOK2* track palatal involvement independently of side. Together, these findings suggest that epigenetic variation acts downstream of genetic predisposition to shape cleft phenotypes.

## Introduction

Orofacial clefts (OFCs) are among the most common congenital malformations, affecting approximately 1 in 700 live births worldwide (1). These conditions result from disturbances during embryonic development that lead to incomplete fusion of the upper lip (cleft lip), palate (cleft palate), or both (2). Approximately 70% of cases are classified as nonsyndromic, occurring without additional congenital anomalies (3). OFCs impose significant functional, aesthetic, and psychosocial burdens on affected individuals, requiring multidisciplinary care from infancy through adulthood.

A striking epidemiological feature of unilateral cleft lip is its non-random laterality distribution with left-sided clefts occurring approximately twice as frequently as right-sided clefts, a pattern consistently observed across diverse populations and geographic regions. This 2:1 ratio strongly suggests that cleft laterality is not a random developmental event but reflects underlying biological mechanisms that preferentially affect left-sided craniofacial development. Despite decades of research, the molecular basis of this laterality bias remains largely unexplained.

Monozygotic twins provide an invaluable model for studying the contribution of genetic and environmental factors to birth defects, as they share nearly identical genetic backgrounds (4). Mirror-image traits, occurring in approximately 25% of monozygotic twin pairs, include hair whorls, birthmarks, and structural anomalies expressed on opposite sides of the body midline (5). Monozygotic twins with mirror-image unilateral clefts—where one twin has a left-sided cleft and the other a right-sided cleft—represent a particularly powerful natural experiment for studying laterality determination while controlling for genetic background. Previous whole-genome sequencing studies of mirror twins with OFCs found no discordant genetic variants that could explain the laterality differences (6), suggesting that epigenetic mechanisms may play a crucial role in modifying cleft laterality (7). DNA methylation, a key epigenetic modification involving the addition of methyl groups to cytosine residues at CpG sites, has emerged as a critical factor in OFC pathogenesis (8–11). By affecting transcription factor binding at gene regulatory regions, DNA methylation can modulate gene expression during critical developmental windows (12, 13).

In this study, we present a comprehensive multi-omic analysis of a female monozygotic twin pair with mirror-image nonsyndromic cleft lip, born to a mother with left-sided microform cleft lip. By integrating whole-genome sequencing with genome-wide DNA methylation profiling and conducting three independent replications, we elucidate the molecular mechanisms underlying both the occurrence and the laterality bias of this common birth defect.

## Results

### Whole-Genome Sequencing Identifies Shared Protein-Altering Variants

Analysis of discordant protein-altering variants between the twins identified a heterozygous variant in *KRT6B* present only in the LCL twin. However, its presence in the unaffected father and heterozygous state made it unlikely to explain the laterality difference. A *de novo* splice site variant in *ACLY* was also identified. While no evidence suggests a role for this gene in craniofacial development, the knockout mouse showed early embryonic (stage E7) lethality. Additionally, we identified a protein-altering variant of uncertain significance in *MUC3A* that was maternally-inherited (heterozygous). Based on the *in-silico* prediction and the likely recessive inheritance pattern, we concluded that this heterozygous maternally-transmitted variant was unlikely to be the risk variant.

Consistent with previous reports (14), we did not identify discordant pathogenic variants that could definitively explain the laterality differences. We therefore focused on maternally transmitted variants shared by both twins and their affected mother (Table 1). This approach identified 16 rare protein-altering variants in craniofacial candidate genes (Table 2).

**Table 1:** Number of variants (CADD >15) at each analytical step towards prioritization of pathogenic variants.

| Filter | Number of Variants |
| --- | --- |
| Protein-altering variants with MAF < 1% | 955 (MZ-1); 939 (MZ-2) |
| Maternally inherited protein-altering variants with MAF < 1% | 393 (MZ-1); 416 (MZ-2) |
| Shared maternally inherited protein-altering variants with MAF < 1% | 387 |
| Variants with CADD≥15 | 175 |
| Variants with CADD≥15 + deleterious (SIFT) | 90 |
| Variants with CADD≥15 + damaging (Polyphen2) | 82 |
| Variants with CADD≥15 + deleterious (SIFT) + damaging (Polyphen2) | 59 |
| Variants with CADD≥15 in Craniofacial genes | 16 |
| Variants with CADD≥15 + deleterious (SIFT) and/or damaging (Polyphen2) in craniofacial genes | 7 |

**Table 2:** Maternally-transmitted shared variants with evidence supporting the roles of the genes in craniofacial development.

| # | Chrom:Pos | Ref/Alt | Gene | HGVS p. | SIFT | PolyPhen | CADD | Facial genes | Craniofacial phenotype | Cleft genes (DB) | ACMG cat | Inheritance |
| --- | --- | --- | --- | --- | --- | --- | --- | --- | --- | --- | --- | --- |
| 1 | 7:7452365 | G/C | *COL28A1* | NP_001032852.1:p.Pro488Arg | tolerated | Possibly damaging | 22.9 | Mid forehead | Nil | Nil | -13 (B) | LR |
| 2 | 9:126503192 | C/T | *MVB12B* | NP_258257.1:p.Arg297Cys | tolerated | Probably damaging | 26.4 | Self-reported chin dimples | Nil | Nil | 0 (VUS) | LR |
| 3 | 5:113028959 | C/T | *MCC* | NP_001078846.1:p.Val952Met | deleterious | Probably damaging | 23.8 | Upper lip center | Nil | Nil | 0 (VUS) | VLR |
| 4 | 13:37590506 | C/T | *POSTN* | NP_006466.2:p.Gly103Ser | tolerated | Probably damaging | 24.9 | Nil | Abnormal ameloblast morphology; Abnormal tooth eruption | Nil | 0 (VUS) | VLR |
| 5 | 21:40296147 | C/T | *DSCAM* | NP_001380.2:p.Arg697Gln | tolerated | benign | 20.9 | Nil | Abnormal cranial cavity morphology; dome cranium; small ears | Nil | -11 (B) | VLD |
| 6 | 4:2833015 | C/G | *SH3BP2* | NP_003014.3:p.Ser505Cys | tolerated | benign | 15.58 | Nil | Abnormal craniofacial bone morphology | Nil | -14 (B) | VLR |
| 7 | 15:89291615 | A/C | *FANCI* | NP_001106849:<br>p.Leu631Phe | tolerated | benign | 15.43 | Nil | Abnormal<br>craniofacial<br>bone<br>morphology;<br>Cleft palate | Nil | -21 (B) | VLR |
| 8 | 20:51790193 | A/G | *SALL4* | NP_065169.1:<br>p.Ser764Pro | tolerated | benign | 21 | Nil | Abnormal<br>craniofacial<br>bone<br>morphology;<br>Cleft palate | Nil | -21 (B) | VLD |
| 9 | 16:2100458 | C/T | *PKD1* | NP_001009944:<br>p.Arg3169Gln | tolerated | benign | 16.98 | Nil | Abnormal<br>craniofacial<br>bone<br>morphology;<br>short maxilla | Nil | -2 (LB) | VLD |
| 10 | 16:81044059 | G/A | *ATMIN* | NP_056066.2:<br>p.Val521Ile | tolerated | benign | 21.6 | Nil | Abnormal<br>craniofacial<br>morphology;<br>micrognathia;<br>Thick upper lip | Nil | -12 (B) | VLR |
| 11 | 5:84064787 | T/C | *EDIL3* | NP_005702.3:<br>p.Ile289Val | tolerated | benign | 22.7 | Nil | Abnormal<br>pinna cartilage<br>morphology;<br>floppy ears | Nil | -5 (LB) | LR |
| 12 | 8:17001798 | T/C | *FGF20* | NP_062825.1:<br>p.Ile79Val | deleterious | Possibly<br>damaging | 26.8 | Nil | Abnormal<br>tooth<br>morphology | Nil | -15 (B) | VLD |
| 13 | 8:117835473 | C/T | *EXT1* | NP_000118.2:<br>p.Val379Ile | tolerated | benign | 16.39 | Nil | Cleft of<br>Secondary<br>palate | Yes | 0(VUS) | VLD |
| 14 | 5:37180175 | G/A | *CPLANE1* | NP_075561.3:<br>p.Pro1860Leu | deleterious | benign | 22.6 | Nil | Cleft of upper lip; Cleft palate (Joubert syndrome) | Yes | -7 (B) | VLR |
| 15 | 14:39180971 | C/T | *PNN* | NP_002678.3:<br>p.Ala421Val | tolerated | benign | 17.38 | Nil | Cleft palate | Yes | -3 (LB) | VLR |
| 16 | 15:74722964 | C/T | *CYP1A1* | NP_001306146:<br>p.Gly45Asp | deleterious | Possibly damaging | 22.8 | Nil | Nil | Yes | -14 (B) | LR |

These variants were novel, or rare in population control databases, altered protein sequences, and ranked among the top 1% most deleterious mutations in the human genome (based on CADD scores ≥ 15). Several of these variants were in genes known to be associated with craniofacial development based on evidence from facial gene scan databases (15), mouse genome informatics, and cleft gene databases. Notably, variants in *FANCI, SALL4, EXT1, CPLANE1*, and *PNN* were identified in genes that, when manipulated in mice, resulted in cleft phenotypes (CleftGeneDB; https://bioinfo.uth.edu/CleftGeneDB) (16)

Table 2 shows mutations that have been considered as deleterious, possibly damaging, or probably damaging by at least one prediction tool (*COL28A1, CPLANE1, CYP1A1, MCC, POSTN, FGF20*, and *MVB12B*). The mutations in *COL28A1, CPLANE1, CYP1A1* and *MVB12B* were shared by the twins, the mother and the unaffected brother. The mutations in *MCC, POSTN* and *FGF20* were shared by the three affected individuals (twin girls and mother) and not present in the unaffected father and brother.

Of these potentially pathogenic protein-altering variants in craniofacial genes, only the *FGF20* variant (p.Ile79Val) was predicted by DOMINO to follow a dominant inheritance pattern; thus becoming our main candidate gene.

Protein function analysis showed that the affected isoleucine residue resides within the fibroblast growth factor family domain, which is critical for receptor binding (17). Analysis of human craniofacial epigenomics data revealed regulatory element signatures within *FGF20*, with the highest enhancer peaks observed during Carnegie stages 13-17, corresponding to critical timepoints for primary palate formation (18). Significant *FGF20* expression appeared in craniofacial tissues from CS13 through CS22 (p<0.05), supporting its role in lip and palate development (19).

Our replication analysis with publicly available data whole genome sequencing data from multiethnic triads affected with nonsyndromic cleft lip and palate (20) did not show significant results; however previous genome-wide association study investigating sex as a modifier of variant risk identified a novel cleft risk locus at chromosome 8p22, which is within a topologically associating domain (TAD) containing *FGF20*. That study also demonstrated robust *Fgf20* expression in the developing mouse maxilla between days E10.5 and E13.5 (21).

### In Silico Analyses Reveal Significant FGF20 Transcription in Craniofacial Tissues During Embryonic Development

To further investigate the role of *FGF20* in human craniofacial development, we examined craniofacial regulatory elements signatures within *FGF20* gene and its spatiotemporal expression during embryonic development. Using data from the expanded encyclopedia of DNA elements (ENCODE) (https://www.encodeproject.org), we identified gene regulatory elements signatures within *FGF20* including enhancer-like, promoter-like and CTCF-binding elements in the first exon and intron of the gene. We further analyzed the Human Craniofacial Epigenomics dataset (GSE97752) from the Cotney Lab (https://www.chopcranio.org/human-craniofacial-development) (22) and confirmed the presence of regulatory element signatures within *FGF20*, revealing craniofacial-specific regulatory elements, evidenced by enhancer peaks within the gene. The highest peaks were observed in developing craniofacial tissue structures between Carnegie stages (CS) 13–17, corresponding to approximately 4-6 weeks of embryonic development which are key timepoints for primary palate formation.

To assess FGF20 expression in human craniofacial tissues during embryogenesis, we analyzed bulk and single-cell RNA sequencing (scRNA-seq) data from human craniofacial tissues collected between 4–8 weeks of gestation (GSE197513, phs002008) (19). Significant *FGF20* expression (p<0.05) appeared in craniofacial tissue structures at CS13 (p=0.001) and remained elevated through CS14 (p=0.007), CS15 (p=0.00096), and CS17 (p=0.01), at fold changes >30, >20, >30, and >14, respectively (Figure 1). This spatiotemporal expression pattern correlates with critical stages of craniofacial tissue development coincident with the timing of palatogenesis, thus supporting a role for *FGF20* in lip and palate development.

**Figure 1:**
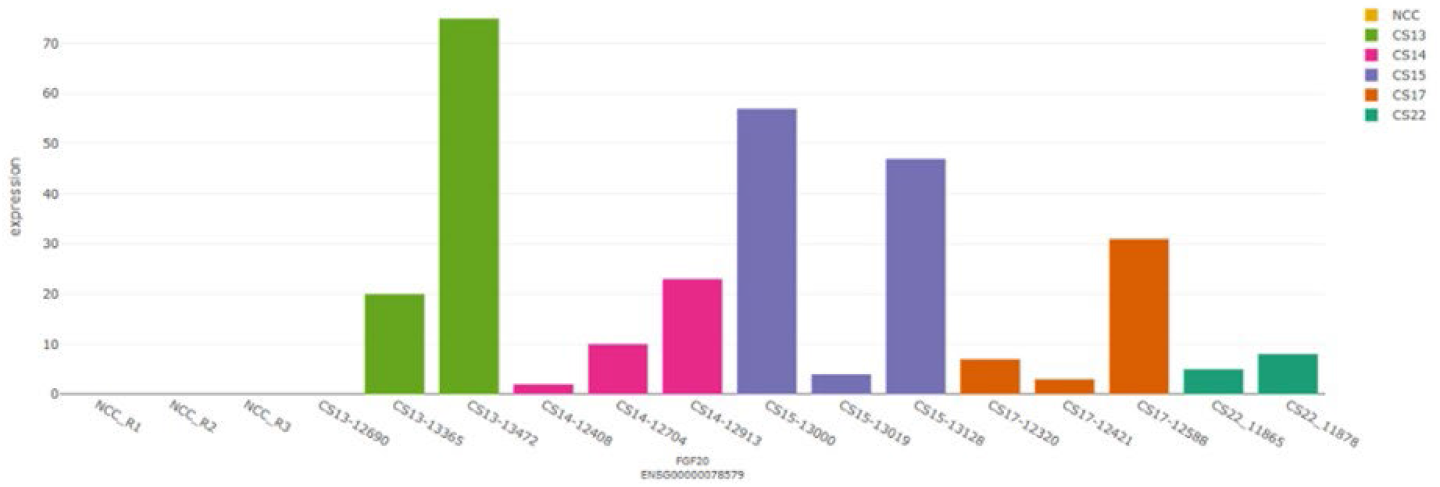
FGF20 expression plot from bulk and single-cell RNA seq of craniofacial tissue between 4-8 weeks post conception. ^*^NCC: Neural Crest Cells; CS: Carnegie Stage.

### DNA Methylation Analysis Identifies Differentially Methylated Regions

Comparison of genome-wide methylation between the mirror twins identified 408 CpG sites with absolute Δβ >5%. Of these, 100 showed higher methylation in the LCL twin, while 308 showed higher methylation in the RCL twin. Enrichment analysis of DMPs within 5kb of gene transcription start sites revealed significant enrichment for the biological process “DNA methylation involved in embryo development” (FDR p^adj^=1.32×10^−13^).

A notable finding was a cluster of differentially methylated CpG sites upstream of the *ZFP57* transcription start site. These sites showed consistently higher methylation in the LCL twin (Δβ=7-12%).

We conducted a replication study of the 408 CpG sites that showed a Δβ ≥ 5% in a small independent cohort. This cohort consisted of individuals with unilateral nonsyndromic CL/P, which were separated based on laterality of the cleft (right: RCL/P or left: LCL/P) and based on source of DNA (Blood or Saliva). Our analysis revealed a borderline significant CpG site in ZFP57 (saliva, p=0.06) and significant CpG sites in *ARID5B* (blood, p=0.002) and *HOOK2* (blood and saliva, p=0.02).

### Additional Replication Confirms Laterality and Cleft Extent-Associated Methylation Patterns

To further investigate these associations (*ZFP57, HOOK2 and ARID5B*), we performed a larger-scale tissue-stratified analysis with an independent replication cohort (n=385), which revealed that methylation differences among cleft subtypes operate along two distinct biological dimensions: cleft laterality (left vs. right side) and cleft extent (CLP vs. CL). Each of the three candidate genes displayed a unique combination of tissue-specific laterality and extent signatures (Tables 3 and 4).

**Table 3.**
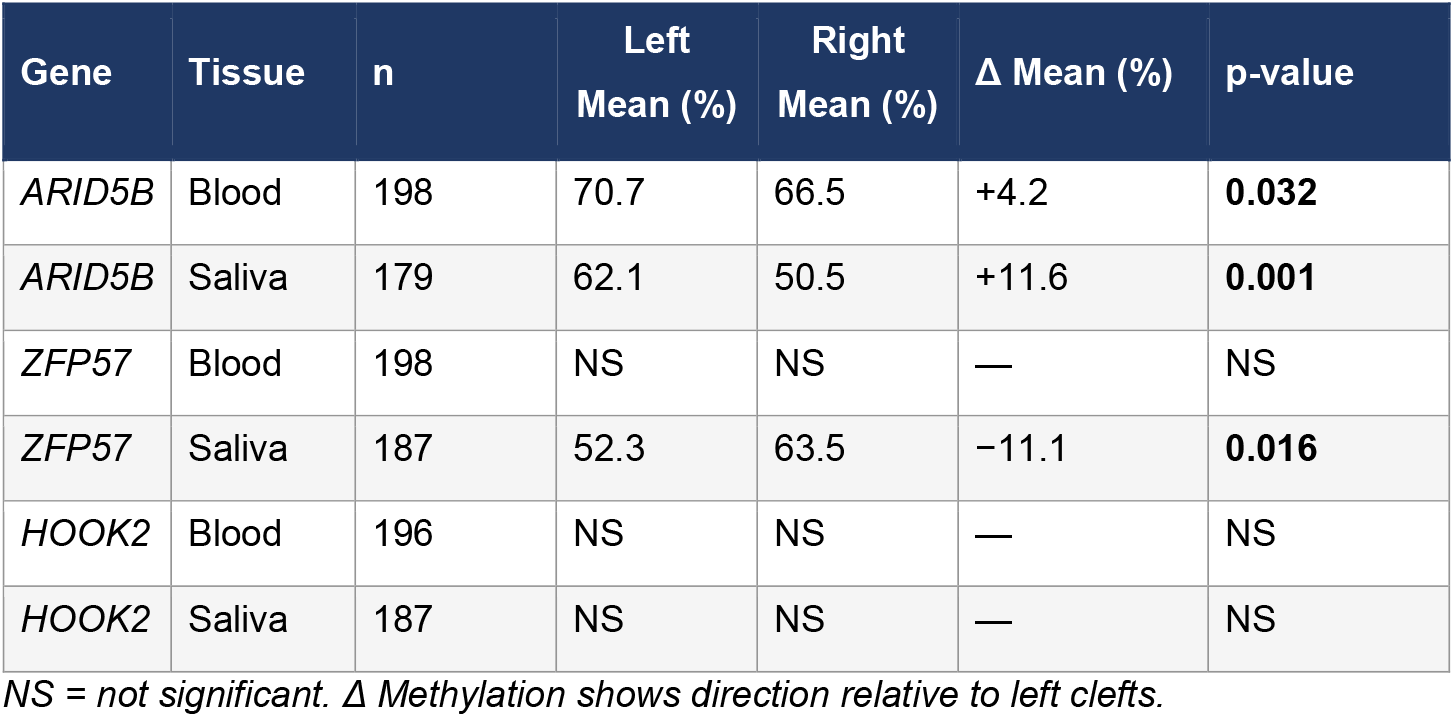
DNA methylation levels by cleft laterality in the replication cohort (n=385)

**Table 4.** Results for four cleft subtypes and comparisons by gene and tissue.

| Gene | Tissue | n | p-value | Median methylation by subtype & significant post-hoc pairs |
| --- | --- | --- | --- | --- |
| *ZFP57* | Blood | 198 | **0.0003\*** | LCLP=56.8%, RCLP=46.1%, LCL=31.0%, RCL=25.4% \| Sig. pairs: LCLP>RCL (p adj=0.0004); LCLP>LCL (p adj=0.019) |
| *ZFP57* | Saliva | 187 | 0.089 | LCLP=40.5%, RCLP=56.2%, LCL=32.9%, RCL=41.4% \| No significant pairs |
| *HOOK2* | Blood | 196 | **<0.0001\*** | LCLP=15.4%, RCLP=18.1%, LCL=19.5%, RCL=20.5% \| Sig. pairs: LCLP<RCL (p adj=0.0003); RCLP<RCL (p adj=0.002); LCLP<LCL (p adj=0.012) |
| *HOOK2* | Saliva | 187 | **0.0008\*** | LCLP=14.5%, RCLP=19.8%, LCL=19.1%, RCL=18.5% \| Sig. pairs: LCLP<LCL (p adj=0.001); LCL>RCL (p adj=0.046) |
| *ARID5B* | Blood | 198 | 0.189 | LCLP=72.5%, RCLP=65.5%, LCL=70.7%, RCL=69.0% \| No significant pairs |
| *ARID5B* | Saliva | 179 | **0.006\*** | LCLP=60.5%, RCLP=54.5%, LCL=70.9%, RCL=44.5% \| Sig. pairs: LCL>RCL (p adj=0.015); LCL>RCLP (p adj=0.024) |

#### ARID5B

The most striking finding emerged from analysis of *ARID5B* (cg22368847), which showed consistently higher methylation in left-sided clefts across both tissue types, representing a cross-tissue, directionally concordant laterality signal. In saliva, left-sided clefts demonstrated mean methylation of 62.1% compared to 50.5% in right clefts (Δ=+11.6%, p=0.001). This laterality pattern was reproduced in blood-derived samples (left: 70.7%, right: 66.5%; Δ=+4.2%; p=0.032), with a smaller but statistically significant effect size consistent with the greater homogeneity of blood-derived methylation (Table 3). In saliva, the four cleft subtypes (H=12.55, p=0.006) revealed that LCL showed the highest median methylation (70.9%), significantly exceeding both RCL (44.5%; p^adj^=0.015) and RCLP (54.5%; p^adj^=0.024) (Table 4). The LCL vs. RCL comparison in saliva (raw p=0.003) represents a laterality-within-CL subtype effect, indicating that the *ARID5B* laterality signal is particularly pronounced in isolated cleft lip cases.

Analysis of human embryonic craniofacial transcriptome data confirmed significant *ARID5B* expression during Carnegie stages 13-22, the precise window of palatal development (Yankee et al., 2023).

#### ZFP57

Analysis of *ZFP57* (cg20228636) revealed a more complex, tissue-divergent pattern. In saliva-derived DNA, right-sided clefts showed significantly higher methylation than left-sided clefts (right: 63.5%, left: 52.3%; Δ=−11.1%; U=3,488, p=0.016) — the opposite direction from *ARID5B*. This directional contrast between genes is notable: sites regulated by the genomic imprinting machinery (*ZFP57*) appear to respond differently to asymmetric craniofacial development signals than chromatin remodeling factors (*ARID5B*).

In blood-derived DNA, *ZFP57* showed no significant laterality effect (U=5,432, p=0.182). Instead, blood revealed a strong and highly significant cleft-extent effect across the four subtypes. CLP subtypes showed markedly higher methylation than CL-only subtypes: median methylation in LCLP (56.8%) was significantly higher than in both LCL (31.0%; p^adj^=0.019) and RCL (25.4%; p^adj^=0.0004) (Table 4). This cleft-extent effect — independent of laterality — suggests that *ZFP57* methylation in blood DNA may reflect the additional epigenetic reprogramming events associated with palatal involvement in CLP, consistent with its role as a master regulator of genomic imprinting at developmental control regions (23–25).

### HOOK2

Analysis of *HOOK2* (cg11738485) revealed no significant laterality effect in either tissue (blood: p=0.282; saliva: p=0.491). Instead, *HOOK2* displayed highly significant cleft-extent effects in *both* types of tissue, the only gene to show cross-tissue consistency for the cleft-extent dimension.

In blood, (p<0.0001), we observed that CLP subtypes were consistently *hypomethylated* relative to CL-only subtypes — a direction opposite to *ZFP57*. LCLP (median=15.4%) was significantly lower than both RCL (median=20.5%; p^adj^=0.0003) and LCL (19.5%; p^adj^=0.012). RCLP (18.1%) was also significantly lower than RCL (p^adj^=0.002) (Table 4).

In saliva, we confirmed that LCLP (median=14.5%) showed significantly lower methylation than LCL (19.1%; p^adj^=0.001). Additionally, a saliva-specific within-CL laterality effect emerged: LCL (mean=32.8%) showed significantly different methylation from RCL (26.5%; p^adj^=0.046), driven by a subset of high-methylation outliers in the LCL group (Table 4).

## Discussion

By leveraging the unique power of monozygotic twins with mirror-image clefts and conducting independent replications — an unbiased genome-wide screen in 38 individuals identifying *ARID5B, ZFP57*, and *HOOK2* as the top candidate loci, followed by a tissue-stratified validation cohort of up to 385 individuals — we have identified gene-specific, tissue-dependent epigenetic signatures that distinguish laterality effects from cleft-severity effects at single-CpG resolution.

Our whole-genome sequencing analysis identified a potentially pathogenic variant in *FGF20* (p.Ile79Val) shared by both twins and their affected mother. Previous studies demonstrated co-expression of *FGF20* and *FGFR2* in developing mouse palatal epithelium (26), and our analysis confirmed significant *FGF20* expression in human craniofacial tissues during critical developmental windows. While this variant likely contributes to cleft susceptibility, it cannot explain the laterality difference since both twins carry the same mutation.

The identification of three genes—*ARID5B, ZFP57*, and *HOOK2*—with reproducible laterality-associated methylation patterns represents a significant advance in understanding cleft etiology. Critically, tissue-stratified analyses reveal that these genes do not simply reflect a single epigenetic dimension but instead capture two distinct biological signals: cleft *laterality* (which side the cleft occurs on) and cleft *extent* (whether the palate is involved).

Understanding this distinction is essential for interpreting the epigenetic architecture of CL/P and for designing future biomarker studies.

***ARID5B*** emerged as the most robustly associated gene across all analyses, showing consistent laterality effects in both blood and saliva — the only gene to do so. The higher methylation in left-sided clefts was particularly pronounced in saliva-derived DNA and in isolated cleft lip cases. As a chromatin remodeling factor and histone demethylase co-activator that regulates transcriptional programs during development (27), *ARID5B* may influence the epigenetic landscape during the asymmetric cellular processes underlying facial development. Its significant expression during Carnegie stages 13-22 positions it precisely at the temporal window when laterality determination and palatal fusion occur (19). The consistency of the *ARID5B* laterality signal across blood and oral epithelial tissues suggests it may reflect a systemic epigenetic asymmetry established during early embryonic patterning rather than a tissue-specific regulatory response.

***ZFP57*** is particularly notable as a master regulator of genomic imprinting that maintains methylation patterns at imprinting control regions during early development (23, 25). Its tissue-divergent profile — right-over-left laterality effect in saliva, and CLP-over-CL extent effect in blood — reveals two distinct regulatory layers at this locus. The saliva-based laterality finding (right>left; Δ=−11.1%, p=0.016) is the opposite in direction from *ARID5B*, suggesting that the genomic imprinting machinery and chromatin remodeling factors respond differentially to the same asymmetric developmental signals. The blood-based cleft-extent effect indicates that palatal involvement is associated with substantially higher *ZFP57* methylation in circulating DNA. Because *ZFP57* regulates imprinting at multiple genomic loci, its differential methylation may have broad downstream effects on imprinted gene expression during craniofacial morphogenesis. Protein interaction analysis confirmed that *ZFP57* and *TFAP2A* participate in networks associated with left-right axis specification, providing a potential mechanistic link between our epigenetic findings and laterality determination (28).

***HOOK2*** displayed the strongest and most consistent cleft-extent epigenetic signal of the three genes, with CLP subtypes significantly hypomethylated relative to CL-only subtypes in both blood and saliva. The LCLP vs. RCL comparison in blood represents the single most statistically significant post-hoc result in the entire replication study. This CLP hypomethylation direction is strikingly opposite to *ZFP57*, suggesting that different molecular mechanisms — imprinting maintenance vs. cytoskeletal gene regulation — respond in opposite directions to palatal involvement. *HOOK2* encodes a microtubule-binding protein critical for centrosome positioning, Golgi ribbon organization, and polarized cell migration (29, 30). During palate development, the mesenchymal cells of the palatal shelves must coordinate cytoskeletal remodeling for shelf elevation and horizontal reorientation — processes that depend heavily on microtubule dynamics and cell polarity (31). Lower *HOOK2* methylation in CLP patients may correspond to altered expression states in these cytoskeletal programs during palatal morphogenesis. Additionally, a saliva-specific LCL vs. RCL difference suggests that *HOOK2* oral epithelial methylation captures an additional laterality-within-CL signal, albeit of borderline significance.

The tissue-specific divergence of epigenetic signals across all three genes underscores a critical methodological consideration for future epigenetic studies of CL/P. Blood and saliva-derived DNA capture fundamentally different aspects of the methylation landscape: blood-derived methylation reflects hematopoietic and systemic developmental programming, while salivary methylation may more directly capture the epigenetic state of craniofacial epithelial progenitors. Consistent with this interpretation, Reinius et al. (2012) demonstrated substantial variation in DNA methylation profiles across purified blood cell lineages, and these tissue-level differences are further amplified in comparisons between blood and mucosal tissues (32, 33). In our data, *ZFP57* exemplifies this divergence most starkly: it shows a laterality effect only in saliva and a cleft-extent effect only in blood, suggesting that distinct regulatory mechanisms operate in parallel compartments. By contrast, *ARID5B*’s laterality signal and *HOOK2*’s extent signal are more broadly shared, consistent with their roles in foundational chromatin and cytoskeletal programs that may be active across multiple tissue lineages.

Our results reveal two distinct epigenetic dimensions in the biology of unilateral CL/P that should be distinguished in future studies: cleft laterality and cleft extent. These dimensions appear to be regulated by different genes and different tissues, and they operate independently. *ARID5B* methylation is primarily a laterality marker; *ZFP57* (blood) and *HOOK2* (both tissues) are primarily cleft-extent markers; and *ZFP57* (saliva) and *HOOK2* (saliva, CL group) bridge both dimensions. This multi-dimensional architecture parallels the known genetic architecture of OFC, in which CLP and CL have partially distinct genetic etiologies and risk loci (2, 9). The epigenetic findings reported here suggest that these genetically distinct developmental programs are also epigenetically distinguishable at specific CpG sites.

The stronger laterality effects observed in isolated cleft lip cases for *ARID5B* and *HOOK2* may reflect differential sensitivity of epigenetic regulation across developmental stages. Cleft lip results from failure of fusion of the maxillary and nasal processes during weeks 4–7 of gestation, while palatal clefting involves secondary palate fusion during weeks 8–12 (2). The presence of palatal involvement (CLP) may introduce additional epigenetic heterogeneity that dilutes the pure laterality signal, explaining why laterality-specific methylation differences are more pronounced in CL-only cases.

A notable demographic difference between the blood and saliva groups in our replication cohort (blood: 92.9% non-White; saliva: 57.5–59.9% non-White across genes) warrants acknowledgment. This population composition difference may contribute to some of the observed tissue divergences and should be explored in future population-stratified analyses. Nonetheless, the directional consistency of findings within each tissue group, and the concordance between our discovery data in the mirror twins and the replication results for *ARID5B* and *ZFP57* laterality effects, support the biological reproducibility of these signals. From a translational perspective, our findings establish the groundwork for developing epigenetic biomarkers for cleft risk assessment and phenotype prediction. *ARID5B* methylation, as the most consistent cross-tissue laterality marker, may have the greatest potential as an accessible biomarker from blood or saliva. The reversible nature of DNA methylation also opens possibilities for preventive interventions, as methylation patterns can be influenced by nutritional factors affecting one-carbon metabolism, including folate, methionine, and choline (13).

Our study has limitations. The discovery analysis was limited to a single twin pair, inherent to the rarity of mirror-image clefts. However, our two-stage replication strategy — an unbiased genome-wide screen followed by a targeted tissue-stratified validation in up to 385 individuals across multiple population groups — substantially strengthens our findings. Population composition differences between tissue groups limit direct cross-tissue comparisons. Future studies should incorporate matched tissue collection (blood and saliva from the same individuals), population-stratified analyses, and mechanistic studies linking the identified CpG sites to gene expression during craniofacial morphogenesis.

## Materials and Methods

### Subjects and Sample Collection

The discovery cohort consisted of a family with female monozygotic twins, their unaffected brother, and their parents. One twin presented with left-sided microform cleft lip (LCL), while the other exhibited right-sided microform cleft lip (RCL). The twins’ mother had a left-sided microform cleft lip; the brother and father had no structural abnormalities (Figure 2). Blood samples were collected from all individuals. The study was approved by the University of Iowa Institutional Review Board (protocols 199804081, 200003065, and 200710721), and all participants provided written informed consent.

**Figure 2:**
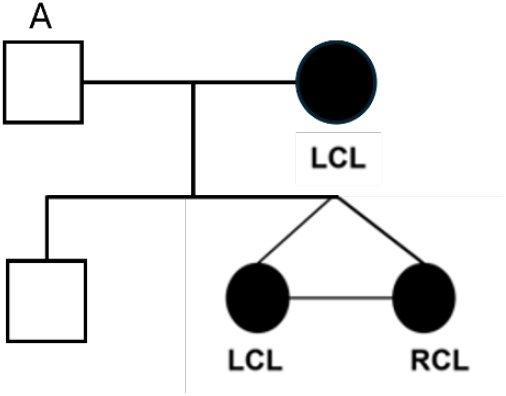
(A) Pedigree showing the inheritance pattern of clefting in the family.

### Whole-Genome Sequencing

Whole-genome sequencing was performed at an average coverage depth of 30×. Sequence data were aligned to the human genome assembly GRCh38 using the Dynamic Read Analysis for GENomics (DRAGEN) pipeline. We prioritized high-confidence variants (genotype quality ≥20, read depth ≥10) that were rare or novel (minor allele frequency <1% in gnomAD). Protein-altering variants (missense and loss-of-function) were assessed using SIFT, PolyPhen2, and CADD. Variants were classified according to the American College of Medical Genetics and Genomics (ACMG) system, and the DOMINO algorithm was used to assess dominant inheritance patterns (34). Our workflow is illustrated and detailed in Figure 3.

**Figure 3:**
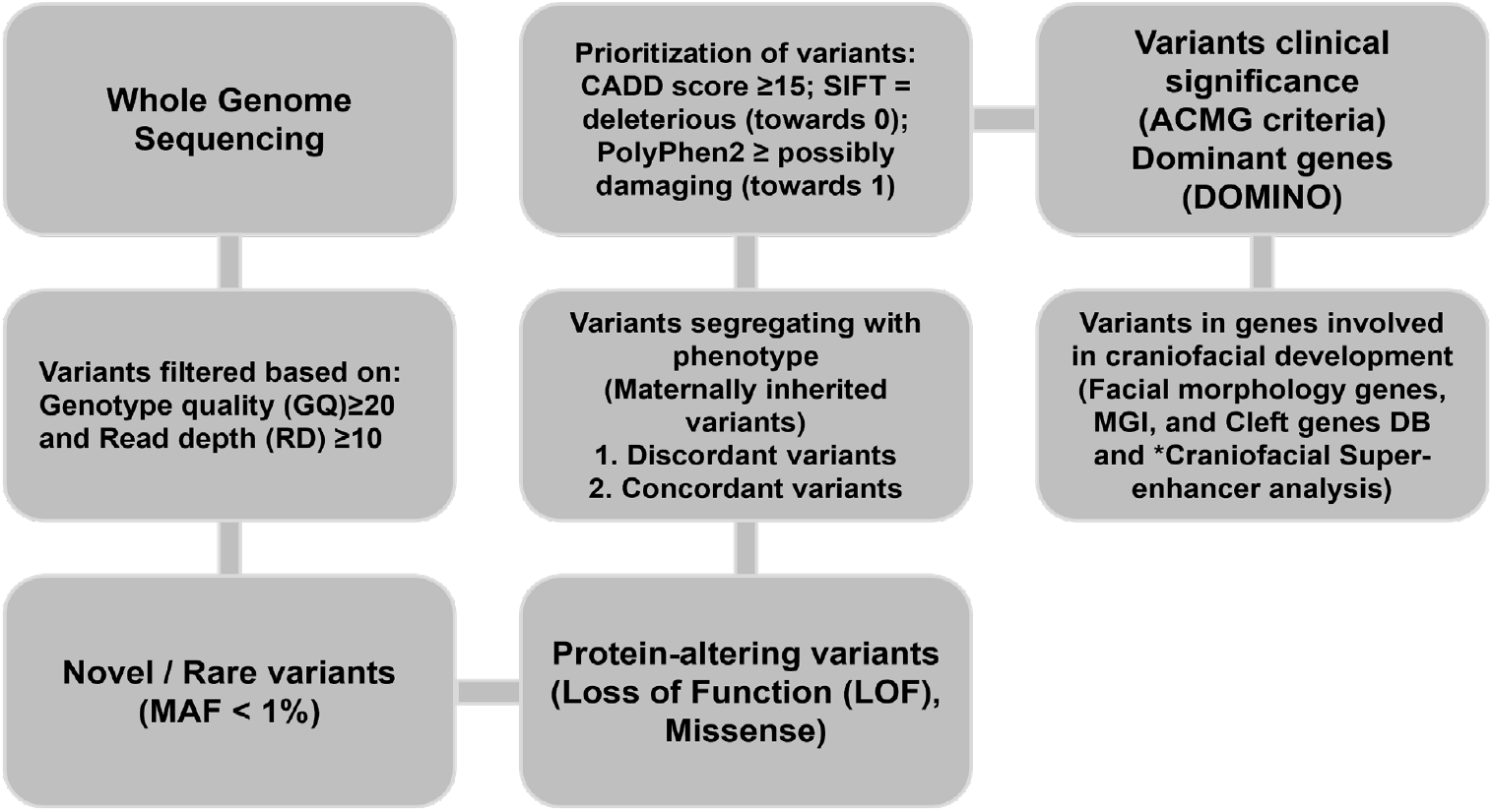
Genomic analytical pipeline used to investigate discordant and concordant pathogenic protein-altering variants in the mirror twins contributing to the risk of cleft.

### Additional Sequencing Analysis

Sanger sequencing confirmed variant segregation in the family for top candidates from Table 2. For replication of the *FGF20* finding, we analyzed the *FGF20* genomic region (full transcriptional span plus 1 kb upstream) in whole-genome sequencing data from 1,611 multiethnic parent-offspring trios with nonsyndromic CL/P (20). Variants were extracted using bcftools and tested for preferential transmission using an allelic transmission disequilibrium test (TDT). A complementary rare variant burden test (MAF <0.1%) was performed using the same TDT framework with gnomAD allele frequencies.

### *In Silico* Functional Analysis

Following the identification of the main candidate genes, we examined craniofacial regulatory elements signatures and their spatiotemporal expression during embryonic development. Using data from the expanded encyclopedia of DNA elements (ENCODE) (https://www.encodeproject.org), we searched for gene regulatory elements signatures including enhancer-like, promoter-like and CTCF-binding elements. We further analyzed the Human Craniofacial Epigenomics dataset (GSE97752) from the Cotney Lab (https://www.chopcranio.org/human-craniofacial-development) (22) to look for presence of regulatory element signatures, revealing craniofacial-specific regulatory elements active during relevant Carnegie stages (CS13-22) (19).

### DNA Methylation Analysis in Twins

Genome-wide DNA methylation profiling was performed using the Illumina EPIC BeadChip array, which evaluates over 850,000 CpG sites. Methylation levels were quantified as β-values (0-1), representing the ratio of methylated to total signal. The EpiDISH package was used for cell-type composition normalization (35). CpG sites with coefficient of variation >20% in gender-matched controls were excluded. Differentially methylated positions (DMPs) with absolute Δβ ≥5% between twins were selected for downstream analysis. The DNA methylation analysis workflow is illustrated in Figure 4.

**Figure 4:**
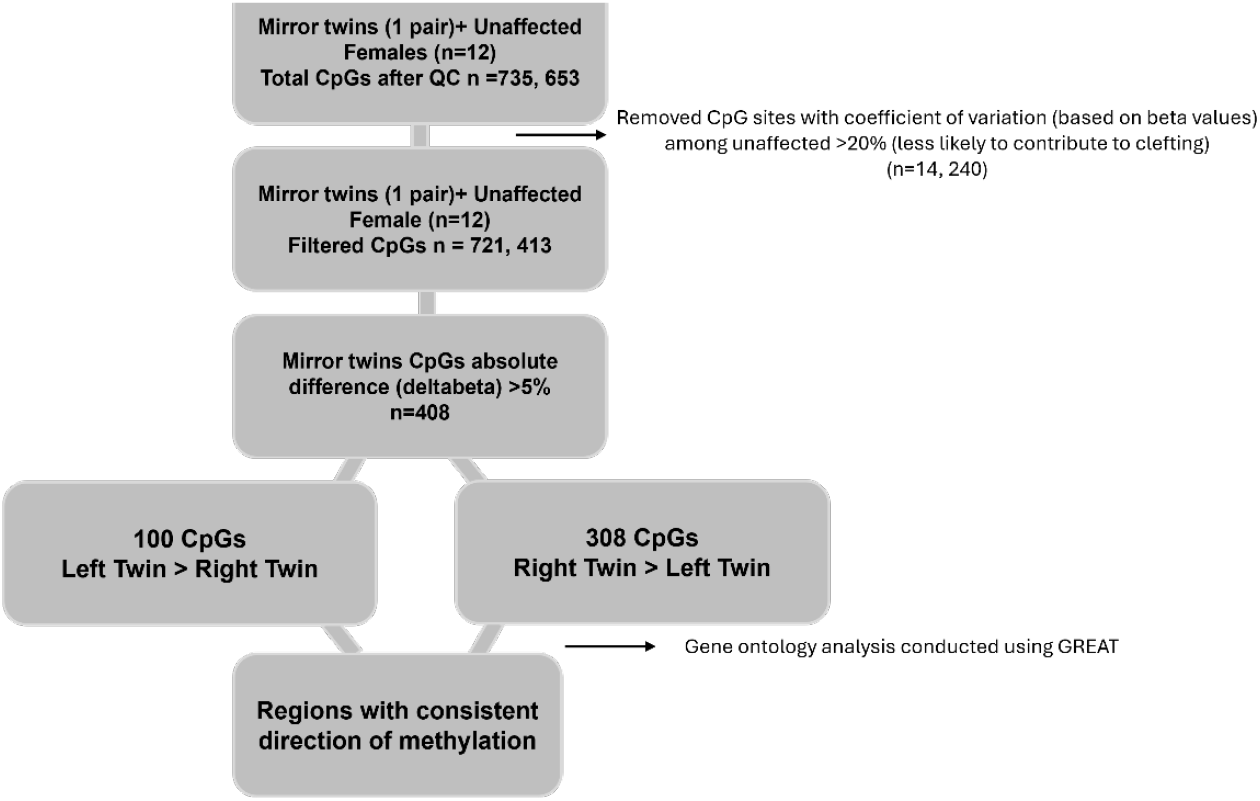
DNA Methylome analytical pipeline.

### Methylation Replication Studies

We performed a ***genome-wide methylation analysis*** (Illumina EPIC array) in 38 individuals with unilateral cleft lip and/or palate, comprising blood samples (right=9, left=13) and saliva samples (right=7, left=9). This screening identified three genes (*ARID5B, ZFP57*, and *HOOK2)* as the top three differentially methylated candidate loci.

We then followed up with a ***targeted methylation analysis of three priority candidate genes*** — *ARID5B* (CpG site cg22368847; n=377), *ZFP57* (CpG site cg20228636; n=385), and *HOOK2* (CpG site cg11738485; n=383) — in individuals with unilateral nonsyndromic CL/P. Each cohort comprised approximately equal proportions of four cleft subtypes: left cleft lip and palate (LCLP), right cleft lip and palate (RCLP), left cleft lip only (LCL), and right cleft lip only (RCL). Critically, analyses were performed separately for blood-derived (*ZFP57*: n=196; *HOOK2*: n=196; *ARID5B*: n=198) and saliva-derived (*ZFP57*: n=187; *HOOK2*: n=187; *ARID5B*: n=179) DNA to account for tissue-specific methylation patterns (32). To compare methylation levels between two groups (e.g., left vs. right clefts), the normality of each group’s distribution was assessed using the Shapiro-Wilk test. The nonparametric Mann-Whitney U test (equivalent to the Wilcoxon rank-sum test) was applied for all two-group comparisons, as the majority of subgroups exhibited significant departures from normality. For comparisons across four cleft subtypes (LCLP, RCLP, LCL, RCL), the Kruskal-Wallis test was used. When the Kruskal-Wallis test indicated a significant difference, pairwise post-hoc comparisons were conducted using Mann-Whitney U tests with Bonferroni correction applied across all six pairwise comparisons (adjusted significance threshold α=0.0083). All analyses were conducted using the SAS® System version 9.4 (SAS Institute Inc., Cary, NC, USA). Statistical significance was set at p<0.05.

## Data Availability

All data produced in the present study are available upon reasonable request to the authors.

## Acknowledgments

We are grateful to all families who voluntarily participated in this study. This work was supported by NIH/NIDCR K01DE027995, University of Iowa College of Dentistry Student Research and the University of Iowa College of Dentistry Clinical/Dental Education Research Initiative Support Program (CRISP).

## Author Contributions

A.L.P., W.A., C.E.S, and L.M-U. contributed to conception and design, data acquisition, analysis and interpretation, and drafted and critically revised the manuscript. C.E.S., L.A.M.P., L.H., P.B., H.K., F.Q., M.C., S.D., J.C.T., A.B., J.C.M., and S.R.V. contributed to conception, data acquisition, analysis, interpretation, and critical revision of the manuscript. All authors gave final approval and agreed to be accountable for all aspects of the work.

## Data Availability Statement

The data supporting the findings of this study are available from the corresponding author upon reasonable request.

## Ethical Approval Statement

The study was approved by the University of Iowa Institutional Review Board and all participants signed informed consents prior to collection of clinical data and biological samples.

## References

1 Mossey, P.A., Little, J., Munger, R.G., Dixon, M.J. and Shaw, W.C. (2009) Cleft lip and palate. Lancet, 374, 1773–1785.

2 Dixon, M.J., Marazita, M.L., Beaty, T.H. and Murray, J.C. (2011) Cleft lip and palate: understanding genetic and environmental influences. Nature Reviews Genetics, 12, 167.

3 Gowans, L.J.J., Al Dhaheri, N., Li, M., Busch, T., Obiri-Yeboah, S., Oti, A.A., Sabbah, D.K., Arthur, F.K.N., Awotoye, W.O., Alade, A.A. et al. (2021) Co-occurrence of orofacial clefts and clubfoot phenotypes in a sub-Saharan African cohort: Whole-exome sequencing implicates multiple syndromes and genes. Molecular genetics & genomic medicine, 9, e1655.

4 Sahu, M. and Prasuna, J.G. (2016) Twin Studies: A Unique Epidemiological Tool. Indian Journal of Community Medicine : Official Publication of Indian Association of Preventive & Social Medicine, 41, 177–182.

5 Stanković-Babić, G., Vujanović, M. and Cekić, S. (2011) Identical twins with “mirror image” anisometropia and esotropia. Srp. Arh. Celok. Lek., 139, 661–665.

6 Takahashi, M., Hosomichi, K., Yamaguchi, T., Nagahama, R., Yoshida, H., Marazita, M.L., Weinberg, S.M., Maki, K. and Tajima, A. (2018) Exploration of genetic factors determining cleft side in a pair of monozygotic twins with mirror-image cleft lip and palate using whole-genome sequencing and comparison of craniofacial morphology. Arch. Oral Biol., 96, 33–38.

7 Wang, L., Liu, Z., Lin, H., Ma, D., Tao, Q. and Liu, F. (2017) Epigenetic regulation of left–right asymmetry by DNA methylation. The EMBO Journal, 36, 2987–2997– 2997.

8 Alvizi, L., Ke, X., Brito, L.A., Seselgyte, R., Moore, G.E., Stanier, P. and Passos-Bueno, M.R. (2017) Differential methylation is associated with non-syndromic cleft lip and palate and contributes to penetrance effects. Scientific reports, 7, 2441.

9 Sharp, G.C., Ho, K., Davies, A., Stergiakouli, E., Humphries, K., McArdle, W., Sandy, J., Davey Smith, G., Lewis, S.J. and Relton, C.L. (2017) Distinct DNA methylation profiles in subtypes of orofacial cleft. Clin. Epigenetics, 9, 63.

10 Xu, Z., Lie, R.T., Wilcox, A.J., Saugstad, O.D. and Taylor, J.A. (2019) A comparison of DNA methylation in newborn blood samples from infants with and without orofacial clefts. Clinical epigenetics, 11, 40.

11 Alvizi, L., Brito, L.A., Kobayashi, G.S., Bischain, B., da Silva, C.B.F., Ramos, S.L.G., Wang, J. and Passos-Bueno, M.R. (2022) mir152 hypomethylation as a mechanism for non-syndromic cleft lip and palate. Epigenetics, in press., 1–18.

12 Boyes, J. and Bird, A. (1991) DNA methylation inhibits transcription indirectly via a methyl-CpG binding protein. Cell, 64, 1123–1134.

13 Garland, M.A., Sun, B., Zhang, S., Reynolds, K., Ji, Y. and Zhou, C.J. (2020) Role of epigenetics and miRNAs in orofacial clefts. Birth defects research, 112, 1635–1659.

14 Takahashi, M., Hosomichi, K., Yamaguchi, T., Nagahama, R., Yoshida, H., Maki, K., Marazita, M.L., Weinberg, S.M. and Tajima, A. (2018) Whole-genome sequencing in a pair of monozygotic twins with discordant cleft lip and palate subtypes. Oral diseases, 24, 1303– 1309.

15 Shaffer, J.R., Orlova, E., Lee, M.K., Leslie, E.J., Raffensperger, Z.D., Heike, C.L., Cunningham, M.L., Hecht, J.T., Kau, C.H., Nidey, N.L. et al. (2016) Genome-Wide Association Study Reveals Multiple Loci Influencing Normal Human Facial Morphology. PLoS Genet, 12, e1006149.

16 Suzuki, A., Jun, G., Abdallah, N., Gajera, M. and Iwata, J. (2018) Gene datasets associated with mouse cleft palate. Data Brief, 18, 655–673.

17 Ohmachi, S., Mikami, T., Konishi, M., Miyake, A. and Itoh, N. (2003) Preferential neurotrophic activity of fibroblast growth factor-20 for dopaminergic neurons through fibroblast growth factor receptor-1c. J. Neurosci. Res., 72, 436–443.

18 Wilderman, A., VanOudenhove, J., Kron, J., Noonan, J.P. and Cotney, J. (2018) High-Resolution Epigenomic Atlas of Human Embryonic Craniofacial Development. Cell reports, 23, 1581–1597.

19 Yankee, T.N., Oh, S., Winchester, E.W., Wilderman, A., Robinson, K., Gordon, T., Rosenfeld, J.A., VanOudenhove, J., Scott, D.A., Leslie, E.J. et al. (2023) Integrative analysis of transcriptome dynamics during human craniofacial development identifies candidate disease genes. Nature Communications, 14, 4623.

20 Mukhopadhyay, N., Bishop, M., Mortillo, M., Chopra, P., Hetmanski, J.B., Taub, M.A., Moreno, L.M., Valencia-Ramirez, L.C., Restrepo, C., Wehby, G.L. et al. (2020) Whole genome sequencing of orofacial cleft trios from the Gabriella Miller Kids First Pediatric Research Consortium identifies a new locus on chromosome 21. Hum. Genet., 139, 215–226.

21 Awotoye, W., Comnick, C., Pendleton, C., Zeng, E., Alade, A., Mossey, P.A., Gowans, L.J.J., Eshete, M.A., Adeyemo, W.L., Naicker, T. et al. (2022) Genome-wide Gene-by-Sex Interaction Studies Identify Novel Nonsyndromic Orofacial Clefts Risk Locus. Journal of dental research, 101, 465–472.

22 Wilderman, A., VanOudenhove, J., Kron, J., Noonan, J.P. and Cotney, J. (2018) High-Resolution Epigenomic Atlas of Human Embryonic Craniofacial Development. Cell Reports, 23, 1581–1597.

23 Li, X., Ito, M., Zhou, F., Youngson, N., Zuo, X., Leder, P. and Ferguson-Smith, A.C. (2008) A Maternal-Zygotic Effect Gene, Zfp57, Maintains Both Maternal and Paternal Imprints. Dev. Cell, 15, 547–557.

24 Mackay, D.J.G., Callaway, J.L.A., Marks, S.M., White, H.E., Acerini, C.L., Boonen, S.E., Dayanikli, P., Firth, H.V., Goodship, J.A., Haemers, A.P. et al. (2008) Hypomethylation of multiple imprinted loci in individuals with transient neonatal diabetes is associated with mutations in ZFP57. Nat. Genet., 40, 949–951.

25 Quenneville, S., Verde, G., Corsinotti, A., Kapopoulou, A., Jakobsson, J., Offner, S., Baglivo, I., Pedone Paolo V., Grimaldi, G., Riccio, A. et al. (2011) In Embryonic Stem Cells, ZFP57/KAP1 Recognize a Methylated Hexanucleotide to Affect Chromatin and DNA Methylation of Imprinting Control Regions. Molecular cell, 44, 361–372.

26 Hosokawa, R., Deng, X., Takamori, K., Xu, X., Urata, M., Bringas, P., Jr. and Chai, Y. (2009) Epithelial-specific requirement of FGFR2 signaling during tooth and palate development. J. Exp. Zool. B Mol. Dev. Evol., 312b, 343–350.

27 Baba, A., Ohtake, F., Okuno, Y., Yokota, K., Okada, M., Imai, Y., Ni, M., Meyer, C.A., Igarashi, K., Kanno, J. et al. (2011) PKA-dependent regulation of the histone lysine demethylase complex PHF2–ARID5B. Nat. Cell Biol., 13, 668–675.

28 de Crozé, N., Maczkowiak, F. and Monsoro-Burq, A.H. (2011) Reiterative AP2a activity controls sequential steps in the neural crest gene regulatory network. Proceedings of the National Academy of Sciences, 108, 155–160.

29 Pallesi-Pocachard, E., Bazellieres, E., Viallat-Lieutaud, A., Delgrossi, M.-H., Barthelemy-Requin, M., Le Bivic, A. and Massey-Harroche, D. (2016) Hook2, a microtubule-binding protein, interacts with Par6α and controls centrosome orientation during polarized cell migration. Scientific reports, 6, 33259.

30 Szebenyi, G., Wigley, W.C., Hall, B., Didier, A., Yu, M., Thomas, P. and Krämer, H. (2007) Hook2 contributes to aggresome formation. BMC Cell Biol., 8, 19.

31 Bush, J.O. and Jiang, R. (2012) Palatogenesis: morphogenetic and molecular mechanisms of secondary palate development. Development, 139, 231–243.

32 Smith, Z.D. and Meissner, A. (2013) DNA methylation: roles in mammalian development. Nature Reviews Genetics, 14, 204–220.

33 Reinius, L.E., Acevedo, N., Joerink, M., Pershagen, G., Dahlén, S.-E., Greco, D., Söderhäll, C., Scheynius, A. and Kere, J. (2012) Differential DNA Methylation in Purified Human Blood Cells: Implications for Cell Lineage and Studies on Disease Susceptibility. PLoS One, 7, e41361.

34 Quinodoz, M., Royer-Bertrand, B., Cisarova, K., Di Gioia, S.A., Superti-Furga, A. and Rivolta, C. (2017) DOMINO: Using Machine Learning to Predict Genes Associated with Dominant Disorders. Am. J. Hum. Genet., 101, 623–629.

35 Teschendorff, A.E., Breeze, C.E., Zheng, S.C. and Beck, S. (2017) A comparison of reference-based algorithms for correcting cell-type heterogeneity in Epigenome-Wide Association Studies. BMC Bioinformatics, 18, 105.

